# Adaptive behavior deficits in individuals with 3q29 deletion syndrome

**DOI:** 10.1101/2023.03.31.23288022

**Authors:** Rebecca M Pollak, T Lindsey Burrell, Joseph F Cubells, Cheryl Klaiman, Melissa M Murphy, Celine A Saulnier, Elaine F Walker, Stormi Pulver White, Jennifer G Mulle

**Affiliations:** Center for Advanced Biotechnology and Medicine, Robert Wood Johnson Medical School, Rutgers University; Department of Pediatrics, School of Medicine, Emory University; Department of Human Genetics, School of Medicine, Emory University; Department of Psychiatry and Behavioral Science, School of Medicine, Emory University; Marcus Autism Center, Children’s Healthcare of Atlanta and Emory University; Neurodevelopmental Assessment & Consulting Services; Department of Psychology, Emory University; Department of Psychiatry, Robert Wood Johnson Medical School, Rutgers University

## Abstract

**Background:** 3q29 deletion syndrome (3q29del) is associated with a significantly increased risk for neurodevelopmental and neuropsychiatric phenotypes. Mild to moderate intellectual disability (ID) is common in this population, and previous work by our team identified substantial deficits in adaptive behavior. However, the full profile of adaptive function in 3q29del has not been described, nor has it been compared to other genomic syndromes associated with elevated risk for neurodevelopmental and neuropsychiatric phenotypes.

**Methods:** Individuals with 3q29del (n=32, 62.5% male) were evaluated using the Vineland Adaptive Behavior Scales, Third Edition, Comprehensive Parent/Caregiver Form (Vineland-3). We explored the relationship between adaptive behavior and cognitive function, executive function, and neurodevelopmental and neuropsychiatric comorbidities in our 3q29del study sample, and we compared subjects with 3q29del to published data on Fragile X syndrome, 22q11.2 deletion syndrome, and 16p11.2 deletion and duplication syndromes.

**Results:** Individuals with 3q29del had global deficits in adaptive behavior that were not driven by specific weaknesses in any given domain. Individual neurodevelopmental and neuropsychiatric diagnoses had a small effect on adaptive behavior, and the cumulative number of comorbid diagnoses was significantly negatively associated with Vineland-3 performance. Both cognitive ability and executive function were significantly associated with adaptive behavior, and executive function was a better predictor of Vineland-3 performance than cognitive ability. Finally, the severity of adaptive behavior deficits in 3q29del was distinct from previously published data on comparable genomic disorders.

**Conclusions:** Individuals with 3q29del have significant deficits in adaptive behavior, affecting all domains assessed by the Vineland-3. Executive function is a better predictor of adaptive behavior than cognitive ability in this population and suggests that interventions targeting executive function may be an effective therapeutic strategy.

## Introduction

3q29 deletion syndrome (3q29del) is a rare (∼1:30,000) (Kendall et al., 2017, Stefansson et al., 2014) genomic disorder associated with substantially increased risk for a variety of neurodevelopmental and neuropsychiatric phenotypes. 3q29del is characterized by a 1.6 Mb typically de novo deletion from the end of the long arm of chromosome 3 (hg19, chr3:195725000-197350000) (Ballif et al., 2008, Glassford et al., 2016, Willatt et al., 2005). The 3q29 deletion confers a 19-fold increased risk for autism spectrum disorder (ASD) (Itsara et al., 2009, Pollak et al., 2019, Sanders et al., 2015), a greater than 40-fold increased risk for schizophrenia (SZ) (Kirov et al., 2012, Marshall et al., 2017, Szatkiewicz et al., 2014, Mulle, 2015, Mulle et al., 2010), and liability for mild to moderate intellectual disability (ID), attention deficit/hyperactivity disorder (ADHD), and anxiety disorders (Ballif et al., 2008, Cox and Butler, 2015, Girirajan et al., 2012, Glassford et al., 2016, Klaiman et al., 2022, Sanchez Russo et al., 2021, Willatt et al., 2005). Previous work by our team using direct clinical assessment tools confirmed these previously reported neurodevelopmental and neuropsychiatric phenotypes in a cohort of individuals with 3q29del and identified novel deficits in cognitive ability, executive function, and adaptive behavior (Klaiman et al., 2022, Murphy et al., 2018, Sanchez Russo et al., 2021). Adaptive behavior deficits had not previously been formally associated with the 3q29 deletion, and the specific adaptive challenges in this population have not been explored.

Adaptive behaviors are age-appropriate skills that an individual performs with self-sufficiency as part of their day-to-day life (Tassé et al., 2012, Sparrow et al., 2016). As such, deficits in adaptive behavior can have significant adverse impacts on an individual’s quality of life and ability to function independently (Simões et al., 2016, Buntinx and Schalock, 2010). Delays in adaptive behavior are part of the diagnostic criteria for Intellectual Disability (American Psychiatric Association, 2013) and many genetic and genomic disorders with phenotypic similarities to 3q29del are associated with adaptive behavior deficits, including Fragile X syndrome, 22q11.2 deletion syndrome, Angelman syndrome, Prader-Willi syndrome, and 16p11.2 deletion and duplication syndromes (Klaiman et al., 2014, Hatton et al., 2003, Glaser et al., 2003, Dykens et al., 1996, Butcher et al., 2012, Antshel et al., 2005, Debbané et al., 2006, Gothelf et al., 2007, Fine et al., 2005, Peters et al., 2004, Dykens et al., 1992, Qureshi et al., 2014, Owen et al., 2018, Green Snyder et al., 2016, Hudac et al., 2020, Hanson et al., 2015). Adaptive behavior has also been found to be adversely impacted in individuals with idiopathic ASD, with the most pronounced delays in socialization skills (Fenton et al., 2003, Yang et al., 2016, Bölte and Poustka, 2002, Carter et al., 1998, Volkmar et al., 1987, Kanne et al., 2011). In ASD, adaptive skills tend to fall significantly below age and cognitive expectations, particularly in individuals without ID (Klin et al., 2007, Kanne et al., 2011, Alvares et al., 2020), and these delays are strongly associated with poor adult outcome (Howlin et al., 2014, Alvares et al., 2020). Because the adaptive behavior profile in 3q29del has not been defined, it is currently unknown whether the specific adaptive behavior deficits in this population are similar to or different from those described in other disorders, and recommendations for clinical care have not been established.

In the present study, we define the profile of adaptive behavior in individuals with 3q29del. We also explore the relationship between adaptive behavior and cognitive ability, executive function, and neurodevelopmental and neuropsychiatric phenotypes. This study is a valuable contribution to our current understanding of 3q29del; adaptive behavior in this population was a previously undefined phenotype with substantial impacts on quality of life. Describing the spectrum of adaptive ability in 3q29del is not only important for understanding the phenotype of the disorder, but also for designing and implementing the interventions and support structures that will enable individuals with 3q29del to thrive. The results from this study will help to guide future research into possible interventions to improve adaptive function in this population.

## Methods

### Study participants

Individuals with 3q29del were recruited from the online 3q29 registry (3q29deletion.org) for 2 days of in-person deep phenotyping, as previously described (Klaiman et al., 2022, Murphy et al., 2018, Sanchez Russo et al., 2021). 32 individuals with 3q29del (62.5% male) were included in the present study. Study participants ranged in age from 4.9-39.1 years (mean = 14.5 ± 8.3 years). See Table 1 for a description of the study sample. This study was approved by Emory University’s Institutional Review Board (IRB00064133) and Rutgers University’s Institutional Review Board (PRO2021001360).

**Table 1.** Demographic information and clinical diagnoses for study participants with 3q29 deletion syndrome (n = 32). Anxiety disorders included generalized anxiety disorder, specific phobia, separation anxiety, and social anxiety disorder.

| | | Mean ± SD | Range |
| --- | --- | --- | --- |
| Age (years) | | 14.50 ± 8.26 | 4.85 - 39.12 |
| Composite IQ | | 73.03 ± 14.18 | 40 - 99 |
| Global Executive Composite | | 67.81 ± 10.68 | 45 - 88 |
|  |  | Percent | N |
| Sex | Male | 62.50% | 20 |
| Race | White | 90.63% | 29 |
|  | Other | 9.37% | 3 |
| Ethnicity | Hispanic/Latino | 3.13% | 1 |
|  | Not Hispanic/Latino | 96.87% | 31 |
| ADHD | Yes | 62.50% | 20 |
| ASD | Yes | 37.50% | 12 |
| Anxiety | Yes | 40.63% | 13 |
| Prodrome/psychosis | Yes | 29.17% | 7 |
| Intellectual disability | Yes | 34.38% | 11 |
| Graphomotor weakness | Yes | 78.13% | 25 |
| Enuresis | Yes | 21.88% | 7 |
| Total number of psychiatric diagnoses | 0 | 3.13% | 1 |
|  | 1 | 6.25% | 2 |
|  | 2 | 28.13% | 9 |
|  | 3 | 34.38% | 11 |
|  | 4 | 15.63% | 5 |
|  | 5 or more | 12.50% | 4 |

### Measures

The measures used in this study were as previously described (Klaiman et al., 2022, Murphy et al., 2018, Sanchez Russo et al., 2021). Briefly, adaptive behavior was assessed using the Vineland Adaptive Behavior Scales, Third Edition, Comprehensive Parent/Caregiver Form (Vineland-3) (Sparrow et al., 2016). In the present study, the Comprehensive Parent/Caregiver Form was completed by the study participant’s parent or guardian via the publisher’s online application (Pearson q-Global). Vineland-3 items are rated based on the frequency with which the individual performs each skill, with higher numbers reflecting stronger adaptive behavior. All study participants received a standard score for the Adaptive Behavior Composite (ABC), as well as for the Communication, Socialization, and Daily Living Skills (DLS) domains. Study participants between 3 and 9 years of age (n = 12) also completed the Motor Skills domain. Cognitive ability was evaluated using the Differential Ability Scales, Second Edition (DAS-II) (Elliott et al., 1990) for individuals under 18 years of age (n = 24) or the Wechsler Abbreviated Scale of Intelligence, Second Edition (WASI-II) (Wechsler, 1999) for individuals 18 years of age and older (n = 8). Executive function was evaluated using the Behavior Rating Inventory of Executive Function, 2^nd^ Edition (BRIEF-2) for participants 18 years of age and younger (n = 26) or the Behavior Rating Inventory of Executive Function for Adults (BRIEF-A) for participants over 18 years of age (n = 6) (Gioia et al., 2015, Roth and Gioia, 2005). The BRIEF-2 and the BRIEF-A ask the informant to rate the study participant’s behaviors associated with nine domains of executive function. Diagnoses of neurodevelopmental and neuropsychiatric phenotypes were reached using gold-standard evaluations and clinician best estimate diagnosis. Additional detail regarding the assessments used in the present study have been previously described (Murphy et al., 2018, Sanchez Russo et al., 2021) and are summarized in the Supplemental Information.

### Analysis

All analyses were performed in R version 4.0.4 (R Core Team, 2008). Due to small sample size, analyses were exploratory and unadjusted p values were reported. Standardized scores were used for the Vineland-3 rather than age equivalents because the cognitive impairment in our study subjects with 3q29del was not severe. Using standardized scores also facilitated comparison of 3q29del scores to published data on other genomic syndromes. Standardized scores were used for measures of cognitive ability and executive function. Statistical analysis was performed using simple linear models implemented using the stats R package (R Core Team, 2008). All models adjusted for age and sex. Comparison of the adaptive behavior profile in 3q29del to other disorders was performed using one-sample t-tests implemented using the stats R package (R Core Team, 2008). Data visualization was performed using the plotly R package (Sievert et al., 2017).

## Results

### Vineland-3 performance in 3q29del

Standardized scores on the Vineland-3 are normally distributed, with a mean of 100 and a standard deviation of 15. On average, participants with 3q29del scored substantially lower than the population mean on the Vineland-3 ABC (3q29del mean = 73.91 ± 13.68; Figure 1). Of the four domain scores, individuals with 3q29del had relatively stronger performance on the Motor Skills domain (3q29del mean = 77.58 ± 16.23; Figure 1). Participants with 3q29del experience the most severe deficit in the DLS domain, with mean scores approaching the clinical cutoff of two standard deviations below the population mean (3q29del mean = 72.50 ± 18.08; Figure 1). Study participants scored approximately 1.5-1.75 standard deviations below the population mean on the Communication and Socialization domains (Communication = 74.16 ± 15.42, Socialization = 75.66 ± 17.34; Figure 1).

**Figure 1.**
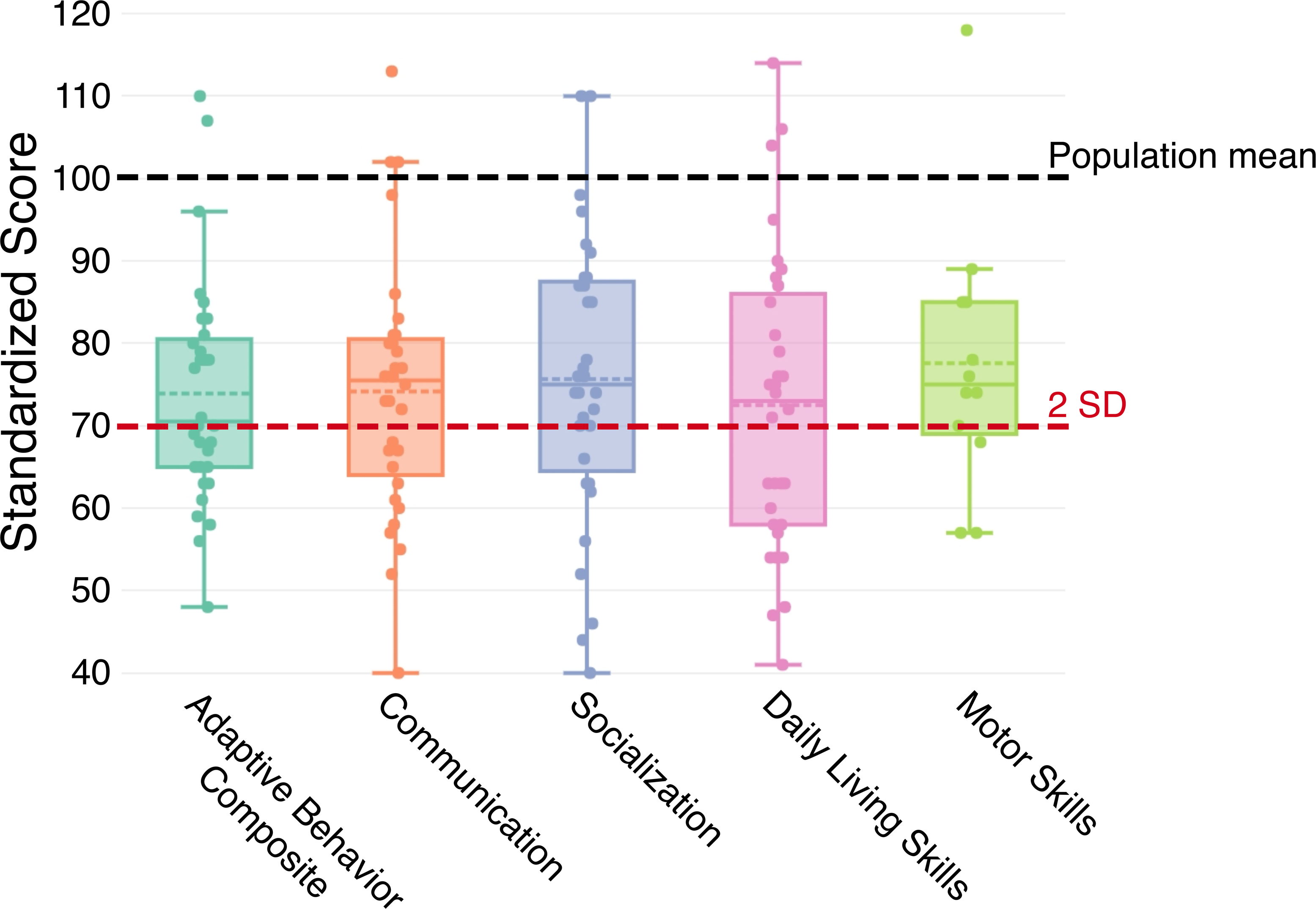
Distribution of scores on the Vineland-3 Adaptive Behavior Composite, Communication domain, Socialization domain, Daily Living Skills domain, and Motor Skills domain for study participants with 3q29del. The dashed black line indicates the population mean of 100, and the dashed red line indicates the clinical cutoff of two standard deviations below the population mean.

To determine whether the domain-level deficits in adaptive behavior are driven by specific challenges versus global deficits, we examined the Vineland-3 subdomains. The subdomains are scored on a v-scale with a mean of 15 and a standard deviation of 3. Within each domain, participants with 3q29del performed relatively consistently across subdomains, with no specific deficits apparent (Figure S1). On average, all subdomain scores were lower than the population mean of 15. Within the Communication domain, individuals with 3q29del had relatively stronger performance on the Expressive Communication subdomain (3q29del mean = 11.72 ± 2.57; Figure S1A) and the most severe deficit on the Written Communication subdomain (3q29del mean = 9.63 ± 3.82; Figure S1A), with an intermediate deficit on the Receptive Communication subdomain (3q29del mean = 10.97 ± 3.35; Figure S1A). Within the Socialization domain, individuals with 3q29del had relatively stronger performance on the Play and Leisure subdomain (3q29del mean = 11.25 ± 3.28; Figure S1B), and comparable deficits on the Interpersonal Relationships and Coping Skills subdomains (Interpersonal Relationships = 10.34 ± 3.50, Coping Skills = 10.84 ± 3.47; Figure S1B). Within the DLS domain, individuals with 3q29del scored consistently across all subdomains (Personal = 10.28 ± 3.80, Domestic = 10.06 ± 3.72, Community = 10.25 ± 4.04; Figure S1C). Within the Motor Skills domain, participants with 3q29del scored consistently across both subdomains, with slightly better performance on the Gross Motor subdomain (3q29del mean = 11.08 ± 3.48; Figure S1D) compared to the Fine Motor subdomain (3q29del mean = 10.83 ± 3.19; Figure S1D). Together, these data demonstrate significant deficits in adaptive behavior in individuals with 3q29del. Further, deficits in adaptive behavior are due to global challenges, rather than severe deficits in specific areas.

### Vineland-3 performance is associated with increasing number of comorbid diagnoses

The 3q29 deletion is associated with substantially elevated risk for ASD and SZ (Kirov et al., 2012, Marshall et al., 2017, Szatkiewicz et al., 2014, Mulle, 2015, Mulle et al., 2010, Itsara et al., 2009, Pollak et al., 2019, Sanders et al., 2015), and individuals with 3q29del commonly have mild to moderate ID (Ballif et al., 2008, Cox and Butler, 2015, Willatt et al., 2005, Girirajan et al., 2012, Glassford et al., 2016, Klaiman et al., 2022, Sanchez Russo et al., 2021). Prior work by our team identified previously unreported associations between the 3q29 deletion and graphomotor weakness and ADHD, as well as a staggering degree of neurodevelopmental and neuropsychiatric comorbidity in this population (Klaiman et al., 2022, Murphy et al., 2018, Sanchez Russo et al., 2021). To understand the relationship between adaptive behavior and neurodevelopmental and neuropsychiatric diagnoses, we compared the ABC score between individuals with 3q29del with and without several neurodevelopmental and neuropsychiatric diagnoses common in this population (Figure 2A). The specific diagnoses examined were ASD, ID, graphomotor weakness, anxiety, prodrome/psychosis, enuresis, and ADHD. On average, individuals with a given neurodevelopmental or neuropsychiatric diagnosis scored lower on the ABC than individuals without the diagnosis, with the exception of enuresis (Figure 2A). Individuals with 3q29del and ID had significantly lower ABC scores than individuals with 3q29del and no ID (p = 0.005); there were no significant differences for any other diagnoses.

**Figure 2.**
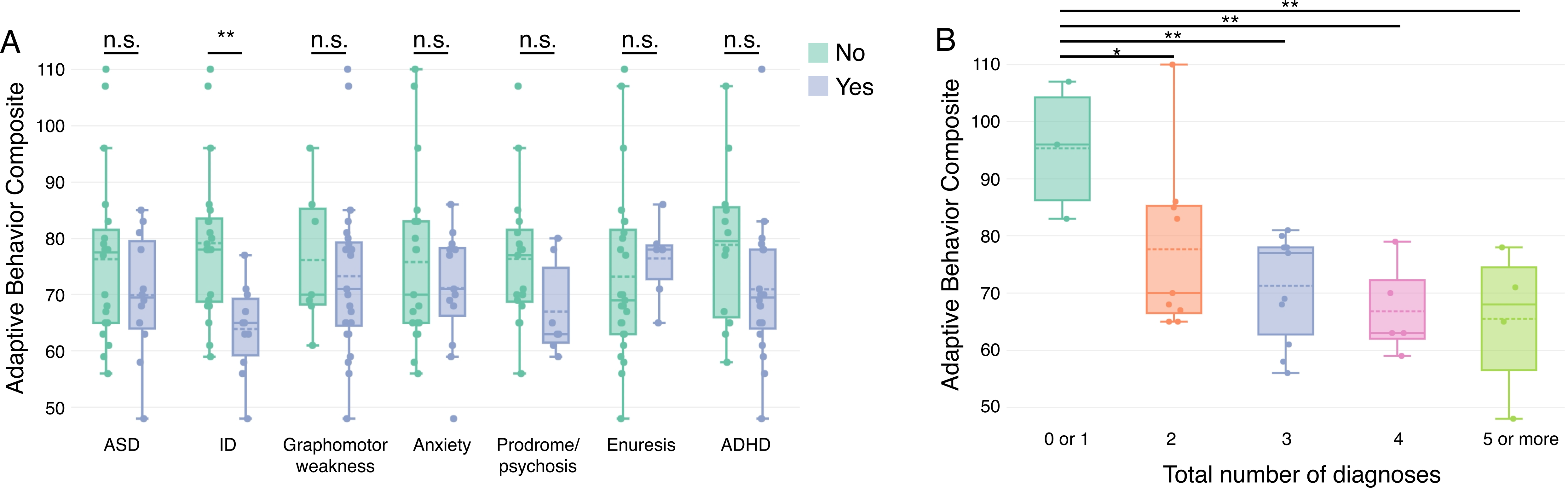
A) Distribution of scores on the Adaptive Behavior Composite for study participants with 3q29del with and without neurodevelopmental and neuropsychiatric diagnoses of interest. B) Adaptive Behavior Composite scores, showing that as the number of an individual’s comorbid neurodevelopmental and neuropsychiatric diagnoses increases, Adaptive Behavior Composite score decreases. n.s., not significant; *, p < 0.05; **, p < 0.001

Due to the extraordinary neurodevelopmental and neuropsychiatric comorbidity associated with the 3q29 deletion, we explored whether the degree of comorbidity was associated with adaptive behavior. We examined comorbidity among the diagnoses named above: ASD, ID, graphomotor weakness, anxiety, prodrome/psychosis, enuresis, and ADHD. Comorbidity in our study population varied, with one individual with zero comorbid diagnoses and four individuals with five or more diagnoses (Table 1, Figure 2B). Over half of our study participants had two (n= 9, 28.13%) or three (n = 11, 34.38%) diagnoses. The average ABC score significantly decreased as the number of comorbid diagnoses increased (Figure 2B). Together, these data suggest that while each individual diagnosis may have a small impact on adaptive behavior, the cumulative effect of multiple neurodevelopmental and neuropsychiatric diagnoses is a far stronger risk factor for impaired adaptive function.

### Executive function is a better predictor of adaptive behavior than cognitive ability in 3q29del

Given that deficits in adaptive function are one of the diagnostic criteria for ID, adaptive behavior and cognitive ability are typically correlated. As expected, individuals with 3q29del and ID had significantly lower adaptive function than individuals with 3q29del and no ID (Figure 2A). We sought to further explore this relationship by examining the association of adaptive behavior and measures of cognitive ability. We found a significant positive correlation between ABC score and composite IQ (r^2^ = 0.162, p = 0.023; Figure 3A). We also identified a significant positive correlation between the Vineland-3 ABC score and verbal IQ (r^2^ = 0.349, p = 0.001; Figure 3B). Nonverbal IQ and spatial ability as measured by the DAS-II were not associated with ABC score (Figure 3C-D). We also examined the association between the Vineland-3 domain scores and cognitive ability. Scores on the Communication domain were significantly positively associated with verbal IQ (r^2^ = 0.311, p = 0.003), but not composite IQ, nonverbal IQ, or spatial ability (Figure S2A-D). Scores on the Socialization domain were significantly positively associated with composite IQ (r^2^ = 0.231, p = 0.049) and verbal IQ (r^2^ = 0.441, p = 0.0004), but not nonverbal IQ or spatial ability (Figure S2E-H). Scores on the DLS domain were significantly positively associated with composite IQ (r^2^ = 0.182, p = 0.007), verbal IQ (r^2^ = 0.291, p = 0.001), and nonverbal IQ (r^2^ = 0.149, p = 0.013), but not spatial ability (Figure S2I-L). Together these data show that adaptive behavior and cognitive ability are positively correlated in individuals with 3q29del.

**Figure 3.**
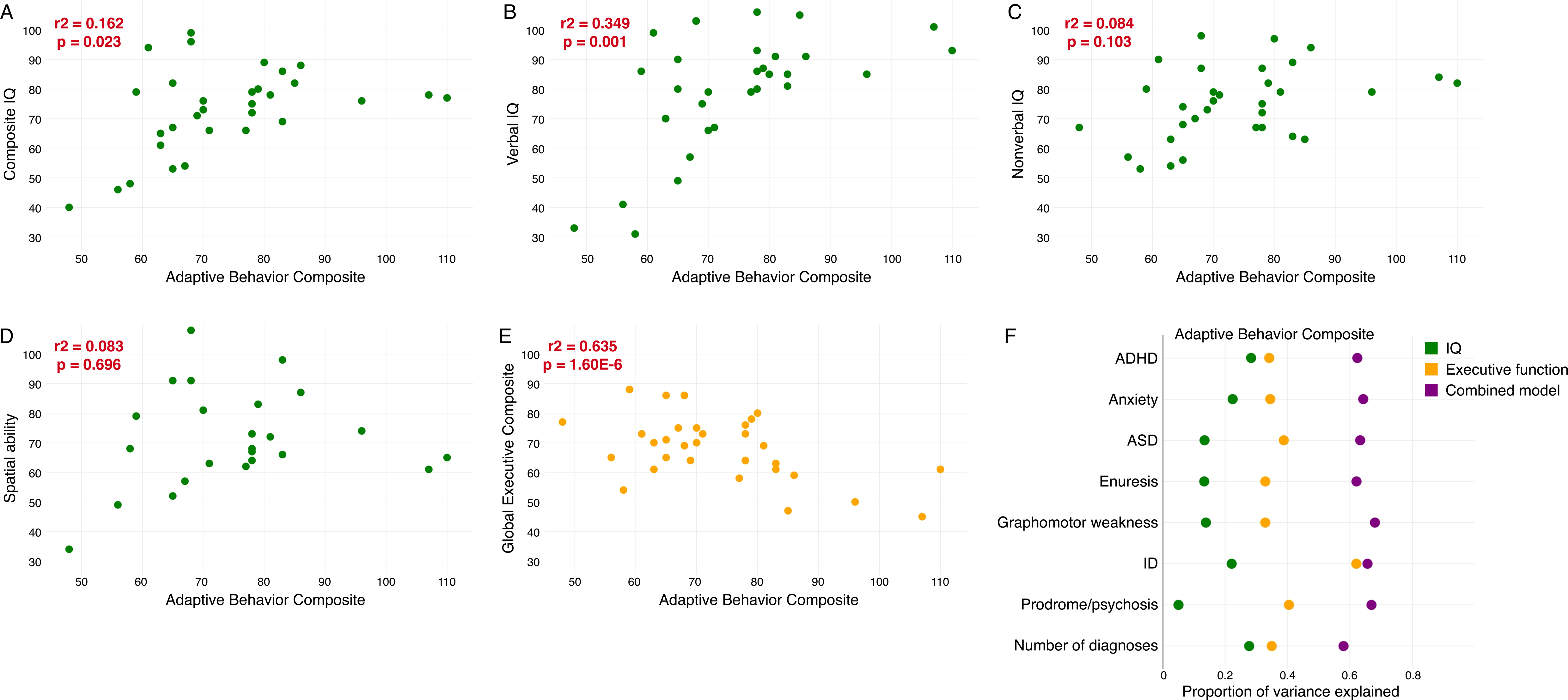
A) Relationship between Adaptive Behavior Composite score and composite IQ score, showing a significant positive correlation. B) Relationship between Adaptive Behavior Composite score and verbal IQ score, showing a significant positive correlation. C) Relationship between Adaptive Behavior Composite score and nonverbal IQ score, showing no correlation. D) Relationship between Adaptive Behavior Composite score and spatial ability, showing no correlation. E) Relationship between Adaptive Behavior Composite score and BRIEF Global Executive Composite score, showing a significant negative correlation. F) Proportion of variance explained in the relationship between Adaptive Behavior Composite score and neurodevelopmental and neuropsychiatric diagnoses by composite IQ, Global Executive Composite score, and a combined model, showing that the Global Executive Composite score is a better predictor of Adaptive Behavior Composite than composite IQ.

In addition to mild to moderate ID, prior work by our group found that a majority of individuals with 3q29del have clinically significant executive function deficits (Klaiman et al., 2022, Murphy et al., 2018, Sanchez Russo et al., 2021). A substantial body of work has shown that executive function is also correlated with adaptive behavior, especially in individuals with ASD (Pugliese et al., 2015, Bertollo and Yerys, 2019, Pugliese et al., 2016, Gilotty et al., 2002). In light of the significant executive function deficits and high rate of ASD in individuals with 3q29del, we explored the association between adaptive behavior and executive function in our study population. It is important to note that unlike measures of cognitive ability, higher scores on the BRIEF correspond to more severe deficits in executive function, and lower scores on the BRIEF correspond to less severe deficits. We found a significant negative correlation between ABC score and the Global Executive Composite score from the BRIEF (r^2^ = 0.635, p = 1.60E-6; Figure 3E). This significant negative correlation held true for all domain scores: Communication (r^2^ = 0.706, p = 9.68E-8; Figure S3A), Socialization (r^2^ = 0.500, p = 0.0004; Figure S3B), and DLS (r^2^ = 0.499, p = 0.0002; Figure S3C). These data highlight the significant relationship between adaptive behavior and executive function in individuals with 3q29del, where individuals with better adaptive function also tend to have better-preserved executive function. Further, we found that the magnitude of the correlations was stronger between adaptive behavior and executive function as compared to those between adaptive behavior and cognitive ability, suggesting that executive function may be a better predictor of adaptive behavior within this population.

We next aimed to test whether cognitive ability or executive function explained more of the variance in adaptive behavior across the different neurodevelopmental and neuropsychiatric diagnoses. For each diagnosis, as well as for the cumulative number of diagnoses, we fit three regression models: one model adjusting for composite IQ, one model adjusting for the BRIEF Global Executive Composite, and one model adjusting for both composite IQ and the Global Executive Composite. We then compared the proportion of variance in the data explained by each model. As expected, we found that the combined model explained the most variance in ABC score for each diagnosis (Figure 3F). Between the models adjusting for either composite IQ or Global Executive Composite, we found that the models adjusting for Global Executive Composite explained more of the variance in ABC score across all diagnosis groups (Figure 3F). We observed a similar trend for the Communication and Socialization domain scores, with the combined model explaining the most variance in score and the Global Executive Composite model outperforming the composite IQ model (Figure S4A- B). The model performance was more variable for the DLS domain score; the combined model explained the most variance in score for all diagnosis groups, but the performance of the Global Executive Composite and composite IQ models varied between diagnoses (Figure S4C). Together, these data reinforce our finding that executive function is a better predictor of adaptive behavior in individuals with 3q29del and suggest that executive function may mediate the impact of neurodevelopmental and neuropsychiatric diagnoses on adaptive behavior.

### Adaptive behavior severity in 3q29del is distinct from other genomic disorders

Adaptive behavior deficits have been identified in multiple genomic disorders with phenotypic similarities to 3q29del, including Fragile X syndrome, 22q11.2 deletion syndrome, and 16p11.2 deletion and duplication syndromes (Klaiman et al., 2014, Hatton et al., 2003, Glaser et al., 2003, Dykens et al., 1996, Butcher et al., 2012, Antshel et al., 2005, Debbané et al., 2006, Gothelf et al., 2007, Fine et al., 2005, Qureshi et al., 2014, Owen et al., 2018, Green Snyder et al., 2016, Hudac et al., 2020, Hanson et al., 2015). We sought to determine whether the profile of adaptive behavior deficits in 3q29del is similar or different from the deficits observed in these other syndromes. We constructed means for each syndrome for the ABC, Communication domain, Socialization domain, and DLS domain, and compared the scores from our cohort of individuals with 3q29del to each syndrome (Table 2). We found that individuals with 3q29del had significantly better performance on the ABC and across all three domains as compared to published data on individuals with Fragile X syndrome (Glaser et al., 2003), indicating that adaptive behavior is better preserved in 3q29del than in Fragile X syndrome. Individuals with 3q29del had significantly higher scores on the ABC and Communication domain as compared to data on 22q11.2 deletion syndrome (Butcher et al., 2012, Antshel et al., 2005, Debbané et al., 2006, Gothelf et al., 2007, Fine et al., 2005), but there was no significant difference on the Socialization or DLS domains. Individuals with 3q29del performed significantly worse on the ABC and all three domain scales as compared to published data on 16p11.2 duplication syndrome (Qureshi et al., 2014, Owen et al., 2018, Green Snyder et al., 2016, Hudac et al., 2020), indicating that individuals with 3q29del have a more substantial deficit in adaptive function than individuals with 16p11.2 duplication syndrome. Finally, individuals with 3q29del had significantly lower scores on the ABC, Socialization domain, and DLS domain as compared to data on 16p11.2 deletion syndrome (Qureshi et al., 2014, Owen et al., 2018, Hudac et al., 2020, Hanson et al., 2015), but there was no significant difference on the Communication domain. Together, these data show that the severity of adaptive behavior deficits in 3q29del is distinct from the deficits reported in other genomic disorders; individuals with 3q29del have better-preserved adaptive function than individuals with Fragile X syndrome or 22q11.2 deletion syndrome, but more severe deficits than those reported in individuals with 16p11.2 deletion or duplication syndromes.

**Table 2.** Cross-disorder comparison between study participants with 3q29 deletion syndrome and published data on Fragile X syndrome (Glaser et al., 2003), 22q11.2 deletion syndrome (Butcher et al., 2012, Antshel et al., 2005, Debbané et al., 2006, Gothelf et al., 2007, Fine et al., 2005), 16p11.2 deletion syndrome (Qureshi et al., 2014, Owen et al., 2018, Hudac et al., 2020, Hanson et al., 2015), and 16p11.2 duplication syndrome (Qureshi et al., 2014, Owen et al., 2018, Green Snyder et al., 2016, Hudac et al., 2020).

|  | 3q29 deletion syndrome |  | Fragile X syndrome |  |  | 22q11.2 deletion syndrome |  |  | 16p11.2 deletion syndrome |  |  | 16p11.2 duplication syndrome |  |  |
| --- | --- | --- | --- | --- | --- | --- | --- | --- | --- | --- | --- | --- | --- | --- |
|  | N | Mean ± SD | N | Mean ± SD | P value | N | Mean ± SD | P value | N | Mean ± SD | P value | N | Mean ± SD | P value |
| Adaptive Behavior Composite | 32 | 73.91 ± 13.68 | 120 | 50.77 ± 20.55 | 9.14E-11 | 238 | 64.25 ± 17.09 | 0.0004 | 277 | 80.18 ± 14.59 | 0.014 | 241 | 81.57 ± 15.11 | 0.003 |
| Communication | 32 | 74.16 ± 15.42 | 120 | 53.46 ± 22.93 | 1.46E-08 | 261 | 66.03 ± 19.54 | 0.006 | 156 | 78.17 ± 12.77 | 0.151 | 133 | 84.04 ± 17.67 | 0.001 |
| Socialization | 32 | 75.66 ± 17.34 | 120 | 61.94 ± 19.52 | 9.62E-05 | 261 | 70.91 ± 18.00 | 0.132 | 156 | 82.17 ± 14.97 | 0.042 | 133 | 85.97 ± 14.99 | 0.002 |
| Daily Living Skills | 32 | 72.50 ± 18.08 | 120 | 47.94 ± 24.19 | 1.15E-08 | 261 | 71.00 ± 20.26 | 0.642 | 81 | 82.20 ± 13.40 | 0.005 | 62 | 81.40 ± 15.40 | 0.009 |

## Discussion

The present study is the first to explore the nuances of adaptive behavior in individuals with 3q29del. We found that individuals with 3q29del have a substantial deficit in adaptive behavior, with mean scores on the ABC and all domains approximately 1.5-1.75 standard deviations below the population mean. However, individual performance is quite variable, with some individuals scoring at or above the population mean, highlighting the high degree of heterogeneity associated with the 3q29 deletion. Notably, the variability in adaptive function observed in our study subjects with 3q29del is substantially less than the variability in adaptive ability in cohorts of individuals with non-syndromic ASD (Klin et al., 2007), suggesting that the adaptive behavior profile in 3q29del is distinct from idiopathic ASD. Adaptive function was significantly worse in individuals with 3q29del and ID as compared to individuals with 3q29del without ID. Further, as the number of comorbid neurodevelopmental and neuropsychiatric diagnoses increased, the mean ABC score decreased, suggesting that it is the degree of comorbidity, rather than a specific diagnosis, that is more strongly associated with adaptive behavior deficits. Both cognitive ability and executive function were significantly correlated with adaptive behavior, but executive function explained more of the variance in adaptive behavior than cognitive ability. This suggests that executive function, not cognitive ability, may be a better predictor of adaptive behavior in this population. Finally, we found that the degree of adaptive behavior deficits in 3q29del is distinct from that reported in Fragile X syndrome, 22q11.2 deletion syndrome, and 16p11.2 deletion and duplication syndromes.

Comorbidity is an important consideration when studying complex disorders such as 3q29del. The traditional medical model silos treatment of different neurodevelopmental and neuropsychiatric disorders with distinct specialists; however, it is possible that examining the comorbidity itself, rather than the individual diagnoses, may provide insight into complex behaviors such as adaptive ability. Indeed, in our study sample we found that the mean ABC score decreased as the number of comorbid diagnoses increased, suggesting that the degree of comorbidity is a predictor of adaptive function. This relationship between comorbidity and adaptive behavior has also been shown in children with ASD, with symptoms of ADHD, anxiety, and depression increasing in severity as adaptive deficits fall farther below cognitive levels (Kraper et al., 2017). In addition, children with ASD and comorbid ADHD have significantly worse adaptive function and quality of life than children with ASD alone (Sikora et al., 2012), highlighting the critical need to understand and appreciate comorbidity as a unique risk factor for adverse outcomes.

Canonically, adaptive behavior and cognitive ability are closely linked; adaptive behavior is a part of the operational definition of ID (Tassé et al., 2012). Because of the well-established association between the 3q29 deletion and mild to moderate ID (Ballif et al., 2008, Cox and Butler, 2015, Klaiman et al., 2022, Sanchez Russo et al., 2021, Willatt et al., 2005), we examined the relationship between adaptive function and measures of cognitive ability. Consistent with the well-established relationship between adaptive behavior and ID, individuals with 3q29del and ID had significantly lower adaptive scores relative to individuals with 3q29del and no ID.

Cognition was positively correlated with adaptive communication, socialization, and daily living skills, demonstrating that individuals with higher IQ tended to also have better-preserved adaptive behavior. Notably, verbal IQ, but not nonverbal IQ or spatial ability, was significantly associated with the overall adaptive composite. This is consistent with prior work by our team suggesting that verbal ability is a driver of overall cognition in individuals with 3q29del (Klaiman et al., 2022). Together, these data confirm an association between adaptive function and cognitive ability in 3q29del.

While adaptive behavior has historically been associated with cognitive ability, a growing body of research is identifying significant correlation between adaptive behavior and executive function. Studies of children with ASD, with and without comorbid ID, have identified a relationship between adaptive behavior and executive function, where higher adaptive behavior is associated with better executive function (Pugliese et al., 2015, Pugliese et al., 2016, Bertollo and Yerys, 2019, Gilotty et al., 2002). Not only do executive functioning deficits predict adaptive deficits in ASD, but they are also associated with comorbidities like anxiety and depression (Udhnani et al., 2020, Wallace et al., 2016). The 3q29 deletion is also associated with ASD and clinically significant executive function deficits (Itsara et al., 2009, Pollak et al., 2019, Sanders et al., 2015, Klaiman et al., 2022, Sanchez Russo et al., 2021). We found a significant correlation between adaptive behavior and executive function in our study subjects with 3q29del, consistent with reports from individuals with idiopathic ASD. We also found that executive function explained more of the variance in adaptive behavior than cognitive ability, suggesting that executive function may be a better predictor of adaptive behavior in this population. Interestingly, this finding has also been reported in a study of children with ASD and low IQ (Bertollo and Yerys, 2019). This relationship between executive function and adaptive behavior may be most relevant in populations with mild to moderate ID, like individuals with 3q29del. Additional studies are needed to further describe the relationship between adaptive behavior, executive function, and cognitive ability in this population.

To put the adaptive behavior deficits in 3q29del into context, we compared the performance by our study subjects to previously published data on individuals with Fragile X syndrome, 22q11.2 deletion syndrome, and 16p11.2 deletion and duplication syndromes. These genomic disorders all have phenotypic similarities to 3q29del: Fragile X syndrome is associated with ID, ASD, and ADHD (Farzin et al., 2006, Hagerman et al., 2017); 22q11.2 deletion syndrome is associated with ID, ASD, ADHD, anxiety, and SZ (McDonald-McGinn et al., 2015, Swillen and McDonald-McGinn, 2015); 16p11.2 deletion syndrome is associated with ID, ASD, and ADHD (Hanson et al., 2015); and 16p11.2 duplication syndrome is associated with ID, ASD, and SZ (D’Angelo et al., 2016). We found that our study subjects with 3q29del performed better than individuals with Fragile X syndrome on all measures; this is not necessarily surprising, given that the cognitive insult in individuals with Fragile X syndrome is much more severe than that present in individuals with 3q29del (Fragile X mean IQ = 56.06 ± 20.35, 3q29del mean IQ = 73.03 ± 14.18, p = 1.40E-7) (Glaser et al., 2003). Study subjects with 3q29del performed significantly worse than individuals with either 16p11.2 deletion or duplication syndromes; IQ was also significantly lower in our study subjects with 3q29del compared to published data on 16p11.2 deletion syndrome (16p11.2 deletion syndrome mean IQ = 83.81 ± 14.81, 3q29del mean IQ = 73.03 ± 14.18, p = 0.0002) (Qureshi et al., 2014, Owen et al., 2018, Hanson et al., 2015) or 16p11.2 duplication syndrome (16p11.2 duplication syndrome mean IQ = 86.87 ± 20.98, 3q29del mean IQ = 73.03 ± 14.18, p = 4.86E-6) (Qureshi et al., 2014, Owen et al., 2018, Green Snyder et al., 2016). The adaptive behavior deficit in our study subjects with 3q29del was most similar to that reported in individuals with 22q11.2 deletion syndrome: individuals with 3q29del performed significantly better than individuals with 22q11.2 deletion syndrome on the ABC and Communication domain, but there was no difference in performance on the Socialization or DLS domains. Additionally, there was no significant difference in IQ between individuals with 3q29del and 22q11.2 deletion syndrome (22q11.2 deletion syndrome mean IQ = 71.25 ± 11.87, 3q29del mean IQ = 73.03 ± 14.18, p = 0.483) (Butcher et al., 2012, Antshel et al., 2005, Debbané et al., 2006, Gothelf et al., 2007). Together, these data suggest that the degree of adaptive behavior deficit in 3q29del is distinct from that observed in similar genomic disorders but may be at least partially driven by differences in cognitive ability between these populations. The similarities and differences in adaptive behavior between 3q29del and these other syndromes is an area ripe with potential for future cross-disorder analysis.

Adaptive behavior assessments like the Vineland-3 measure an individual’s ability to perform age-appropriate tasks (Sparrow et al., 2016). As an individual ages, these tasks become more complex; by adulthood, a person with typical adaptive behavior skills should be able to live and function independently. Thus, deficits in adaptive behavior can be associated with poor adult outcome, particularly in individuals with ASD who have the cognitive capacity to be self-sufficient (Howlin et al., 2014, Alvares et al., 2020), and they can have an adverse effect on quality of life (Simões et al., 2016, Buntinx and Schalock, 2010). Self-sufficiency is critical to quality of life, especially in adulthood; a study of adults with ID found that those adults with better adaptive skills and fewer support needs experienced a higher quality of life (Simões et al., 2016). However, it is important to note that the degree of disability is not an insurmountable hurdle to achieving good quality of life. Rather, quality of life is reached through a combination of an individual’s cognitive and adaptive deficits and the age- and ability-appropriate supports that are provided to them (Buntinx and Schalock, 2010). Thus, it is critical to identify adaptive behavior challenges early, especially in high-risk populations such as individuals with 3q29del, so that early interventions and appropriate supports can be provided to improve long-term outcomes and maximize quality of life.

The high rate of adaptive behavior deficits in 3q29del emphasizes the need for early intervention in this population. Adaptive skills can be targeted through modalities such as occupational therapy to teach age-appropriate motor and daily living skills (Reed and Sanderson, 1999), which is a common component of many early intervention programs. Moreover, we found that the Written Communication subdomain is a relative weakness in individuals with 3q29del, which measures reading and writing skills. This is consistent with prior work by our team identifying significant deficits in visual-motor integration, particularly motor coordination and graphomotor abilities, in this population (Sanchez Russo et al., 2021, Pollak et al., 2022a), highlighting the need for fostering writing skills. Additionally, we have reported that individuals with 3q29del have relatively well-preserved expressive and social communication skills (Pollak et al., 2022b, Pollak et al., 2019); thus, communication-focused interventions should leverage these relative strengths.

Executive function is another modality in this population that could be targeted for treatment. Because executive function is highly correlated with adaptive behavior in individuals with 3q29del, this is an area of therapeutic intervention that could yield substantial improvements in day-to-day function. There have been multiple studies showing that therapeutic coaching can improve executive functioning in older adolescents and young adults (Parker and Boutelle, 2009, Goudreau and Knight, 2018, Franklin and Franklin, 2012). In younger children, integrative mind-body training, a mindfulness-based intervention, has been shown to improve executive function in children as young as 4 years of age (Tang et al., 2012), and the Unstuck and On Target Program is a widely used evidenced-based practice to foster executive functioning skills in children on the autism spectrum (Kenworthy et al., 2014). These data emphasize the malleable nature of executive function across the lifespan; integrating some of these targeted interventions for executive function into early intervention programs for individuals with 3q29del may be an extremely fruitful therapeutic avenue.

While this is the first detailed study of adaptive behavior in individuals with 3q29del, it is not without limitations. First, due to our small sample size our analyses were exploratory. Therefore, it will be critical to replicate our findings in a larger cohort of individuals with 3q29del. We were also unable to assess the effects of race and ethnicity on adaptive function, as our present sample is overwhelmingly white and non-Hispanic. Ongoing efforts to expand study recruitment to more underrepresented minorities will improve our ascertainment, and future studies will ideally have a more representative study sample. Finally, the majority of our study subjects were children (mean age = 14.5 ± 8.3 years); as such, most have not yet passed the age at onset for SZ and related psychotic disorders. It would be beneficial to expand subject recruitment to older adults that have passed the age at onset for these disorders to truly understand the relationship between psychosis and adaptive behavior in this population.

The present study is the first to examine details of adaptive behavior in individuals with 3q29del. We identified significant deficits in global adaptive behavior, as well as in specific areas of communication, socialization, daily living skills, and motor development. The deficits observed were relatively consistent across domains and subdomains, with no specific areas of relative weakness or strength, which is distinct from other neurodevelopmental disorders such as idiopathic ASD. We found that executive function, not cognitive ability, was a better correlate of adaptive behavior in our study subjects; because executive function is a malleable skill, this is a promising area for therapeutic intervention. Based on these data and previous work by our team, we recommend that all individuals with 3q29del should be evaluated for adaptive behavior deficits, so that any challenges can be identified and diagnosed as early as possible. Early diagnosis followed by early intervention in this population is a promising avenue to improved long-term outcomes, quality of life, and ability to function independently.

## Data Availability

The datasets used and/or analyzed during the current study are available from the corresponding author on reasonable request and via NDAR (Collection #2614).

## Acknowledgements

Acknowledgements: We gratefully acknowledge our study population, the 3q29 deletion community, for their participation and commitment to research. This study was funded by the National Institute of Mental Health grant R01 MH110701 (PI Mulle).

Competing interests: CAS reports receiving royalties from Pearson Assessments for the Vineland-3. All other authors report no competing interests.

Ethics approval and consent to participate: This study was approved by Emory University’s Institutional Review Board (IRB00064133) and Rutgers University’s Institutional Review Board (PRO2021001360).

Consent to participate: All study subjects gave informed consent prior to participating in this study.

Consent to publish: Not applicable. Welfare of animals: Not applicable.

Availability of data and materials: The datasets used and/or analyzed during the current study are available from the corresponding author on reasonable request and via NDAR (Collection #2614).

Funding: NIMH R01 MH110701 (PI Mulle)

Author’s contributions: RMP performed the statistical analysis, produced all figures and tables, and wrote the manuscript. TLB, JFC, CK, CAS, EFW, and SPW performed the clinical evaluations of 3q29 deletion study participants. TLB, JFC, CK, MMM, CAS, EFW, and SPW helped with data interpretation. JGM edited the manuscript and provided guidance on analyzing and interpreting data. JGM was the principal investigator responsible for study direction. All authors participated in commenting on the drafts and have read and approved the final manuscript.

## Figure Legends

