## Supplemental Information for "Adaptive behavior deficits in individuals with 3q29 deletion syndrome"

**Supplemental Methods**

*Vineland-3*

The Vineland-3 is a standardized parent interview designed to assess the study participant’s ability to complete day-to-day tasks (Sparrow et al., 2016). The Vineland-3 is a measure of overall adaptive behavior across three domains: Communication, Socialization, and Daily Living Skills. Study participants between 3-9 years of age also were assessed using the Motor Skills domain. The Adaptive Behavior Composite score is calculated from the domain scores and serves as an estimate of the participant’s overall level of adaptive ability.

In the present study, the Vineland-3 Comprehensive Parent/Caregiver form was completed by the study participant’s parent or guardian using the online Pearson q-Global web tool. Scores for analysis were calculated using Pearson q-Global. Age-normed standardized scores were used for analysis. Standardized scores have a mean of 100 and a standard deviation of 15. Higher scores on the Vineland-3 indicate less impaired adaptive behavior.

*DAS-II*

Cognitive ability in study participants less than 18 years of age was assessed using the DAS-II. The DAS-II is comprised of three subtests that measure verbal, nonverbal, and spatial ability. Overall cognitive ability is summarized as a standardized General Conceptual Ability composite score. The DAS-II has two different versions that are administered according to the age of the study participant. The DAS-II Early Years Battery is administered to individuals 6 years of age and younger, and the DAS-II School Age Battery is administered to individuals ages 7 to 18. The DAS-II Early Years Battery has two versions that are administered based on the age and developmental ability of the participant: the Lower Early Years and the Upper Early Years. In the present study, seven participants were administered the DAS-II Early Years Battery (six participants 6 years of age and younger and one 11 year old participant with extremely limited verbal ability), and 17 participants were administered the DAS-III School Age Battery.

In the present study, age-normed standardized scores were used for analysis. Score conversion was performed according to the companion algorithm (Elliott et al., 1990). Standardized scores have a mean of 100 and a standard deviation of 15. Higher scores on the DAS-II indicate less impaired cognitive ability.

*WASI-II*

The WASI-II is a brief measure of cognitive ability appropriate for individuals ages 6 to 90 years (Wechsler, 1999). The WASI-II estimates verbal and nonverbal cognitive ability, and a full-scale IQ is generated to measure overall cognitive ability. In the present study, eight participants 18 years of age or older were assessed using the WASI-II.

Age-normed standardized scores were used for analysis. Score conversion was performed according to the companion algorithm (Wechsler, 1999). Standardized scores have a mean of 100 and a standard deviation of 15. Higher scores on the WASI-II indicate less impaired cognitive ability.

*BRIEF*

The BRIEF is a measure of executive function across nine domains: inhibiting distractions, self-monitoring, shifting, emotional control, initiation, working memory, planning, organization, and task monitoring. For participants 18 years of age or younger, the parent/informant forms (BRIEF-2) were completed by a parent or guardian, and for participants over 18 years of age the Adult Version forms (BRIEF-A) were completed by the participant.

In the present study, the BRIEF was completed by the parent/guardian (BRIEF-2) or participant (BRIEF-A) using the publisher’s website (PARiConnect). Age-normed scores were used for analysis. Scores were calculated using PARiConnect. Higher scores on the BRIEF indicate more impaired executive function.

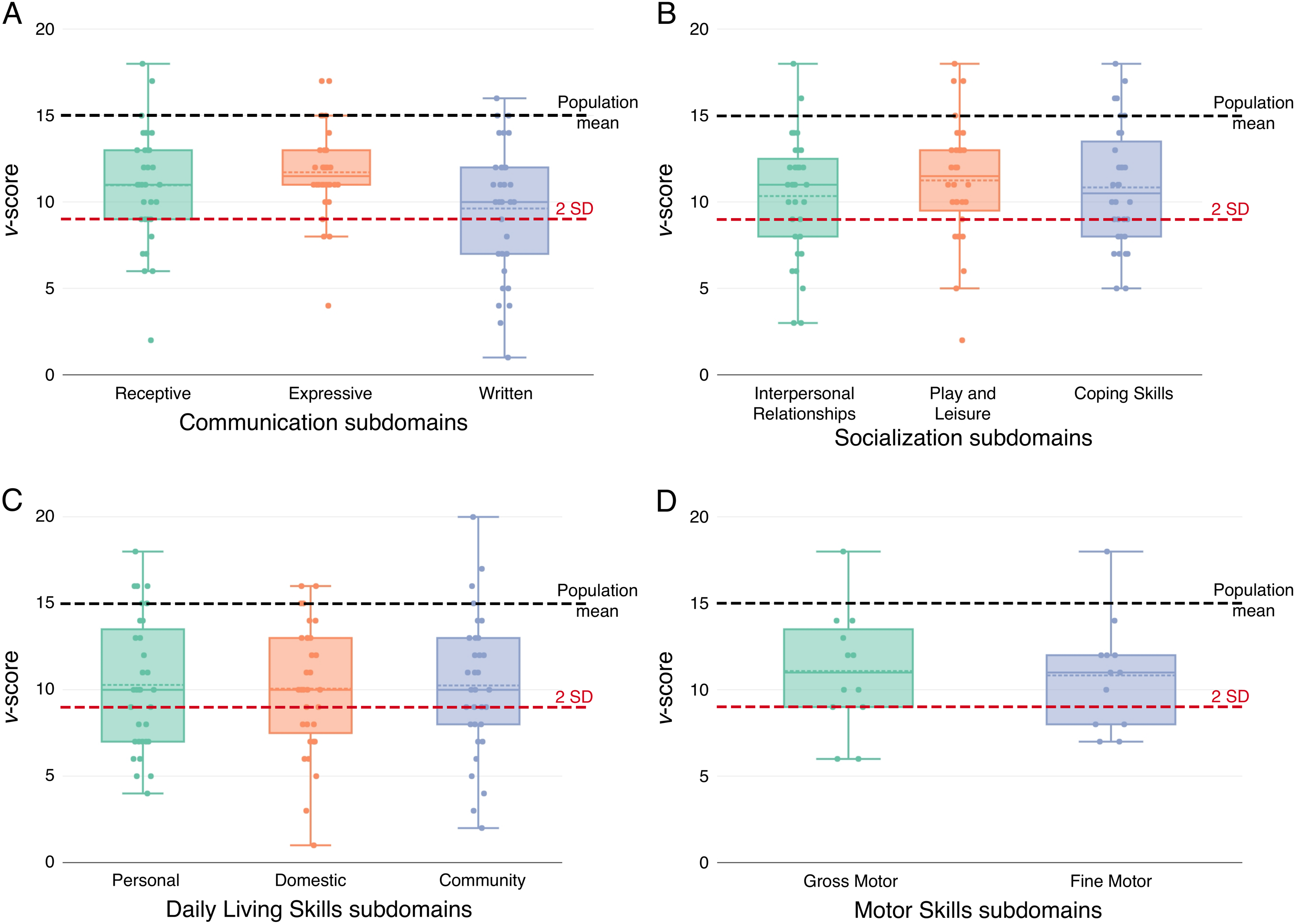

**Figure S1. A)** Scores for study participants with 3q29del on the Communication subdomains: Receptive Communication, Expressive Communication, and Written Communication. **B)** Scores for study participants with 3q29del on the Socialization subdomains: Interpersonal Relationships, Play and Leisure, and Coping Skills. **C)** Scores for study participants with 3q29del on the Daily Living Skills subdomains: Personal, Domestic, and Community. **D)** Scores for study participants with 3q29del on the Motor Skills subdomains: Gross Motor and Fine Motor. The black dashed line indicates the population mean of 15, and the red dashed line indicates the clinical cutoff of two standard deviations below the population mean.

**Figure S2. A)** Relationship between Communication domain scores and composite IQ, showing no correlation. **B)** Relationship between Communication domain scores and verbal IQ, showing a significant positive correlation. **C)** Relationship between Communication domain scores and nonverbal IQ, showing no correlation. **D)** Relationship between Communication domain scores and spatial ability, showing no correlation. **E)** Relationship between Socialization domain scores and composite IQ, showing a significant positive correlation. **F)** Relationship between Socialization domain scores and verbal IQ, showing a significant positive correlation. **G)** Relationship between Socialization domain scores and nonverbal IQ, showing no correlation. **H)** Relationship between Socialization domain scores and spatial ability, showing no correlation. **I)** Relationship between Daily Living Skills domain scores and composite IQ, showing a significant positive correlation. **J)** Relationship between Daily Living Skills domain scores and verbal IQ, showing a significant positive correlation. **K)** Relationship between Daily Living Skills domain scores and nonverbal IQ, showing a significant positive correlation. **L)** Relationship between Daily Living Skills domain scores and spatial ability, showing no correlation.

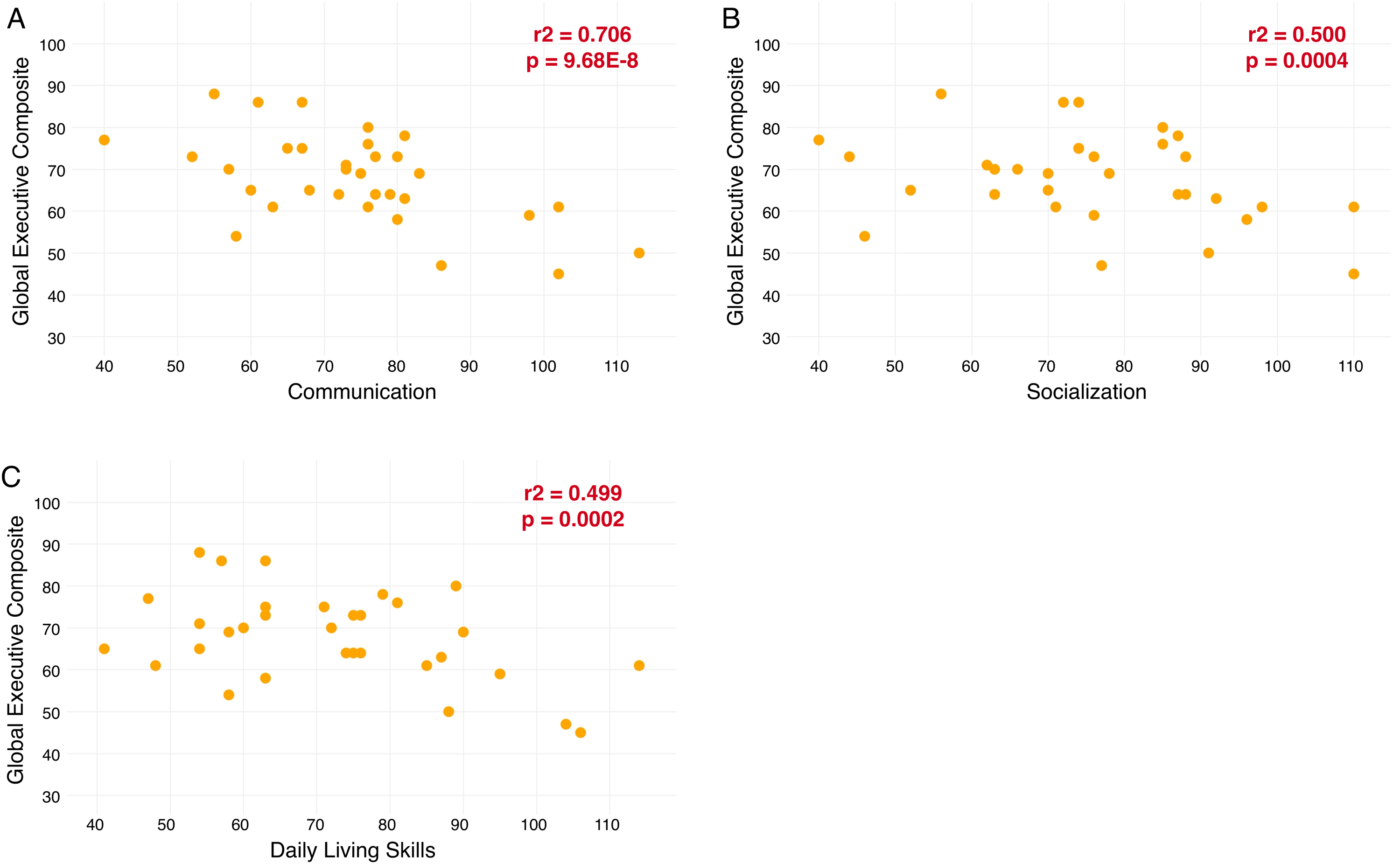

**Figure S3. A)** Relationship between Communication domain scores and BRIEF Global Executive Composite scores, showing a significant negative correlation. **B)** Relationship between Socialization domain scores and BRIEF Global Executive Composite scores, showing a significant negative correlation. **C)** Relationship between Daily Living Skills domain scores and BRIEF Global Executive Composite scores, showing a significant negative correlation.

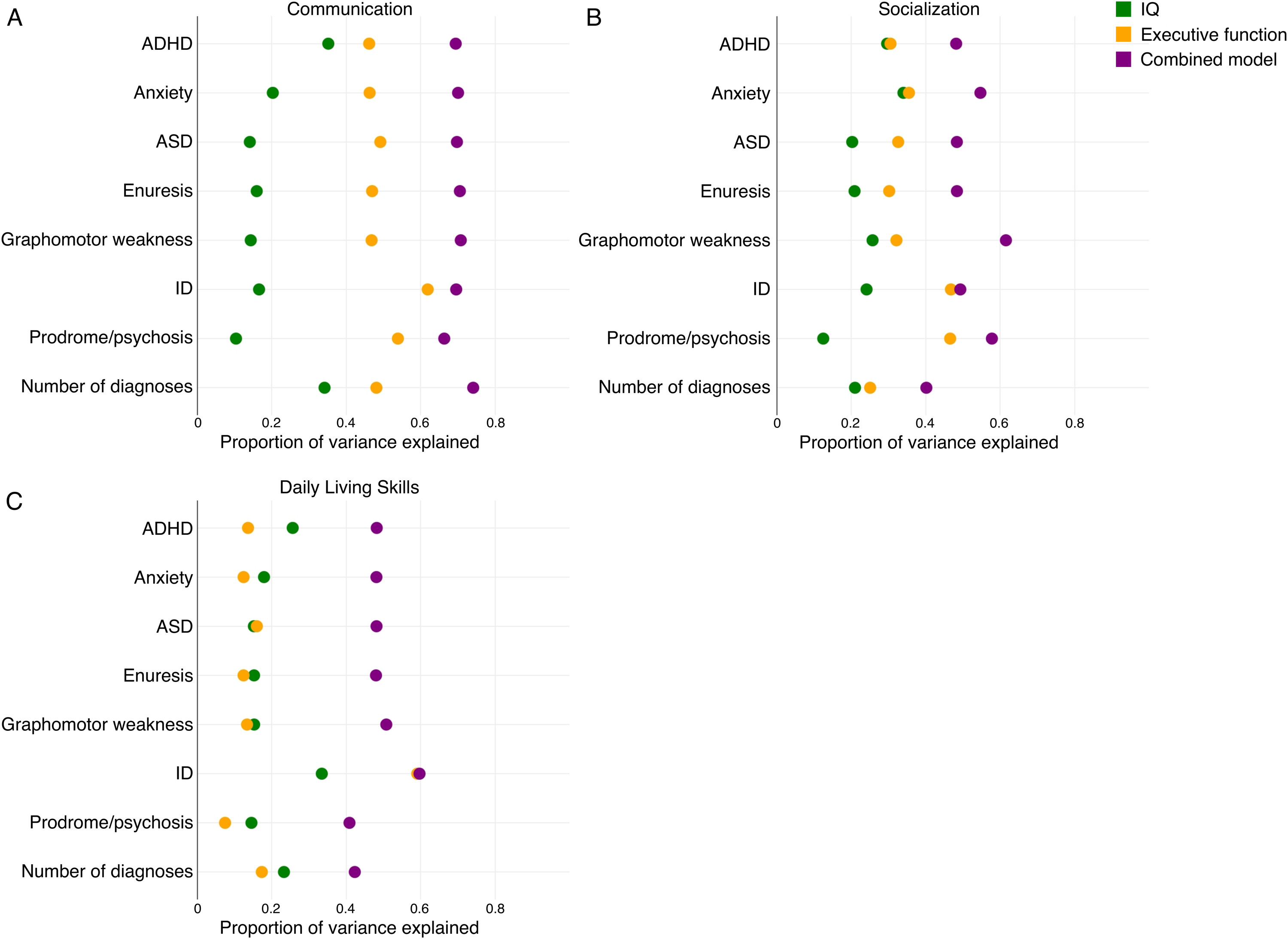

**Figure S4. A)** Proportion of variance explained in the relationship between Communication domain score and neurodevelopmental and neuropsychiatric diagnoses by composite IQ, Global Executive Composite score, and a combined model, showing that the Global Executive Composite score is a better predictor of Communication domain score than composite IQ. **B)** Proportion of variance explained in the relationship between Socialization domain score and neurodevelopmental and neuropsychiatric diagnoses by composite IQ, Global Executive Composite score, and a combined model, showing that the Global Executive Composite score is a better predictor of Socialization domain score than composite IQ. **C)** Proportion of variance explained in the relationship between Daily Living Skills domain score and neurodevelopmental and neuropsychiatric diagnoses by composite IQ, Global Executive Composite score, and a combined model, showing that composite IQ is a better predictor of Daily Living Skills domain score than the Global Executive Composite score.
